# Artificial Intelligence enhanced R1 maps can improve lesion detection in focal epilepsy in children

**DOI:** 10.1101/2025.05.23.25326669

**Authors:** Georgia Doumou, Felice D’Arco, Matteo Figini, Hongxiang Lin, Sara Lorio, Rory Piper, Jonathan O’Muircheartaigh, J Helen Cross, Nikolaus Weiskopf, Daniel C Alexander, David W Carmichael

**Affiliations:** Department of Imaging Physics & Engineering, School of Biomedical Engineering and Imaging Sciences, King’s College London, London, UK; UCL Great Ormond Street Institute for Child Health, London, UK; Centre for Medical Image Computing, Department of Computer Science, University College London, London, UK; Research Center for Healthcare Data Science, Zhejiang Lab, Hangzhou, China; Department for Forensic and Neurodevelopmental Sciences, Institute of Psychiatry, Psychology and Neuroscience & Research Department of Early Life Imaging, School of Biomedical Engineering and Imaging Sciences, & MRC Centre for Neurodevelopmental Disorders, King’s College London, London, UK; Department of Neurophysics, Max Planck Institute for Human Cognitive and Brain Sciences, Leipzig, Germany; Felix Bloch Institute for Solid State Physics, Faculty of Physics and Earth Sciences, Leipzig University, Leipzig, Germany; Wellcome Centre for Human Neuroimaging, Institute of Neurology, University College London, 12 Queen Square, London WC1N 3AR, UK

## Abstract

**Background and purpose:** MRI is critical for the detection of subtle cortical pathology in epilepsy surgery assessment. This can be aided by improved MRI quality and resolution using ultra-high field (7T). But poor access and long scan durations limit widespread use, particularly in a paediatric setting. AI-based learning approaches may provide similar information by enhancing data obtained with conventional MRI (3T). We used a convolutional neural network trained on matched 3T and 7T images to enhance quantitative R1-maps (longitudinal relaxation rate) obtained at 3T in paediatric epilepsy patients and to determine their potential clinical value for lesion identification.

**Materials and Methods:** A 3D U-Net was trained using paired patches from 3T and 7T R1-maps from n=10 healthy volunteers. The trained network was applied to enhance paediatric focal epilepsy 3T R1 images from a different scanner/site (n=17 MRI lesion positive / n=14 MR-negative). Radiological review assessed image quality, as well as lesion identification and visualization of enhanced maps in comparison to the 3T R1-maps without clinical information. Lesion appearance was then compared to 3D-FLAIR.

**Results:** AI enhanced R1 maps were superior in terms of image quality in comparison to the original 3T R1 maps, while preserving and enhancing the visibility of lesions. After exclusion of 5/31 patients (due to movement artefact or incomplete data), lesions were detected in AI Enhanced R1 maps for 14/15 (93%) MR-positive and 4/11 (36%) MR-negative patients.

**Conclusion:** AI enhanced R1 maps improved the visibility of lesions in MR positive patients, as well as providing higher sensitivity in the MR-negative group compared to either the original 3T R1-maps or 3D-FLAIR. This provides promising initial evidence that 3T quantitative maps can outperform conventional 3T imaging via enhancement by an AI model trained on 7T MRI data, without the need for pathology-specific information.

## Introduction

The surgical treatment of focal epilepsy is the only current curative option available and should be considered early in the disease course if medication is ineffective[1]. Identification and accurate delineation of epileptic lesions such as Focal Cortical Dysplasia (FCD) using MRI plays a critical role in surgical evaluation particularly in a paediatric setting[2]. FCDs are often characterized by subtle GM cortical thickness extension and smoothed GM-WM borders not easily seen in conventional MRI[3]. Therefore, a considerable number of patients considered for epilepsy surgery do not have an MRI visible lesion[2] (termed ‘MR negative’) and this is associated with poor surgical outcome[4, 5]. 7T MRI has added value for epilepsy lesion identification and characterisation[6–8] via increased resolution and contrast[6] but has limited availability and is usually characterised by long acquisition times.

One intriguing possibility is the ability of artificial intelligence (AI) in visual learning tasks to enhance the appearance and resolution of MRI data. While AI models have been proposed to generate 7T-like MR images from 3T data, there are few demonstrations of 3T image enhancement in patients with brain abnormalities, and even less that address diagnostic yield. To achieve enhancement a neural network must be trained so that it contains the information necessary to accurately predict the high-resolution appearance of brain structure from low resolution data. Further, it is desirable for the resolution enhancement learnt to be generalisable – i.e. when presented with new data from a source different to that used for training it must retain accuracy of representation. These requirements often necessitate extremely large and varied datasets. Quantitative MRI maps are related to tissue properties regardless of acquisition sequence, hardware or other conditions[9]. This property makes maps highly reproducible across scanners (∼1%[10] that could allow a network to be trained on quantitative MRI maps from one site and applied on new data from other sites with similar fidelity. Of those available, patch-based approaches are particularly interesting for clinical applications as they enable a single matched pair of images to provide many thousands of training examples dramatically reducing the need for large training datasets. Operating at this smaller scale may obviate the need for pathological training examples, where instead diversity across brain structures provides suitable generalisability relying on intensity transformations while operating below the scale of brain morphology.

Therefore, we artificially enhanced R1 (1/T1) maps using CNNs and Image Quality Transfer (IQT), methods that have previously shown promising results in image enhancement[11–17] and assessed their clinical value in focal epilepsy by investigating the following: a) Could the IQT pipeline trained only healthy control data be applied in epilepsy patients with cortical malformations previously detected by clinical MRI (MRI-lesion positive)? b) If so, do the AI enhanced R1 maps increase the visibility of the lesions in comparison to 3T R1 maps or conventional 3T FLAIR images? c) Can the AI enhanced R1 maps improve the sensitivity of lesion detection by aiding the identification of potential cortical abnormalities in clinical 3T MRI lesion negative cases?

## Materials and Methods

Our strategy consists of three main steps: 1) Train a Convolutional Neural Network (CNN) variant based on a previously optimised network (classical 3D isotropic U-Net)[16,18,19] on healthy controls scanned both at 3T and 7T using established qMRI protocols[10, 20]. Healthy control subjects were used as they were able to tolerate the long 7T scan times. 2) Use the trained CNN network to enhance 3T data from a paediatric cohort with focal epilepsy including both MRI positive and negative cases, 3) Assess the image quality and clinical radiological value.

### IQT Network Training

#### Data acquisition

Quantitative R1 maps were obtained in n=10 healthy subjects (mean age 28 ± 4 years) at both 3T (1mm isotropic resolution) and 7T (0.5mm isotropic resolution) at the by Max Planck Institute for Human Cognitive and Brain Sciences, Leipzig, Germany (MPI-CBS) (DataMPI) using multi-parameter mapping (MPM) protocol based on 3D multi-echo GRE sequence acquisitions with variable flip angles[10,20,21].

#### Image processing

Images were processed with the hMRI toolbox[22] to generate 3T and 7T R1 maps. The 1mm resolution 3T R1 maps were coregistered and resliced (4th degree b-spline interpolation) to match the 0.5mm 7T resolution in SPM12. Brain masks were generated and applied to restrict the volume for training and reconstruction reducing computational requirements.

#### IQT network and training

The U-|Net was implemented in Python 3 using Keras library 2.3.1 (www.keras.io) with Tensorflow 1.5 (www.tensorflow.org) backend on a Nvidia Tesla V100 (32 GB). The 3T/7T R1 maps from each subject was used to create a library of paired 3T-to-7T patches using a sliding window to extract 32x32x32 patches with steps of 8x8x4 resulting in ∼36500 patches in total. The patches with a percentage of >80% background voxels were excluded. Remaining patches were used to train the 3D U-Net[16,19]. The network was trained repeatedly where 9 subjects were used to train and a reconstruction was performed on the remaining unseen dataset to test its accuracy. The following parameters were used for all experiments following Lin et al[23]; ADAM (learning rate = 0.0001) was used as the optimizer and the pixel-wise mean squared error (MSE) as the loss function. For the convolutions (and up convolutions) 4 filters were used with 2x2x2 kernel size. An early stopping was set when the validation loss didn’t decrease after 10 epochs. Mean total training time was approximately 51 hours (epoch time ∼ 3 hours), convergence was obtained after 14 epochs and validation MSE loss was 0.11.

### IQT Application in paediatric focal epilepsy

#### Data acquisition

The paediatric focal epilepsy group recruited consisted of n=33 (mean age 15±4 years) patients being evaluated for surgery at Great Ormond Street Hospital, UK (*Table 1* & *Figure 1*). MPM data were incomplete for subjects 16 and 23 and therefore they were excluded from further analysis. The remaining n=31 subjects consisted of two cohorts; MR positives, where a lesion had been visually detected based on radiological review of images from a 3T epilepsy protocol (n=17) and MR negatives, where no lesion was visually detected (n=14) at the time of the MPM acquisition. One subject had a repeated scan (referred to as subjects 3 and 8). T Both scans were reconstructed and included for image quality analysis, but only one of them (subject 8) was used in the clinical evaluation. The following subjects were excluded from the lesion identification (see *Figure 1*); Subject 14 (severe motion artefacts); Subjects 1, 21 and 24 (incomplete electro-clinical data); Subject 15 (later diagnosed with genetic generalized epilepsy). Subject 27 was excluded from the comparison with 3D-FLAIR (3D-FLAIR was not retrievable).

**Fig. 1.**
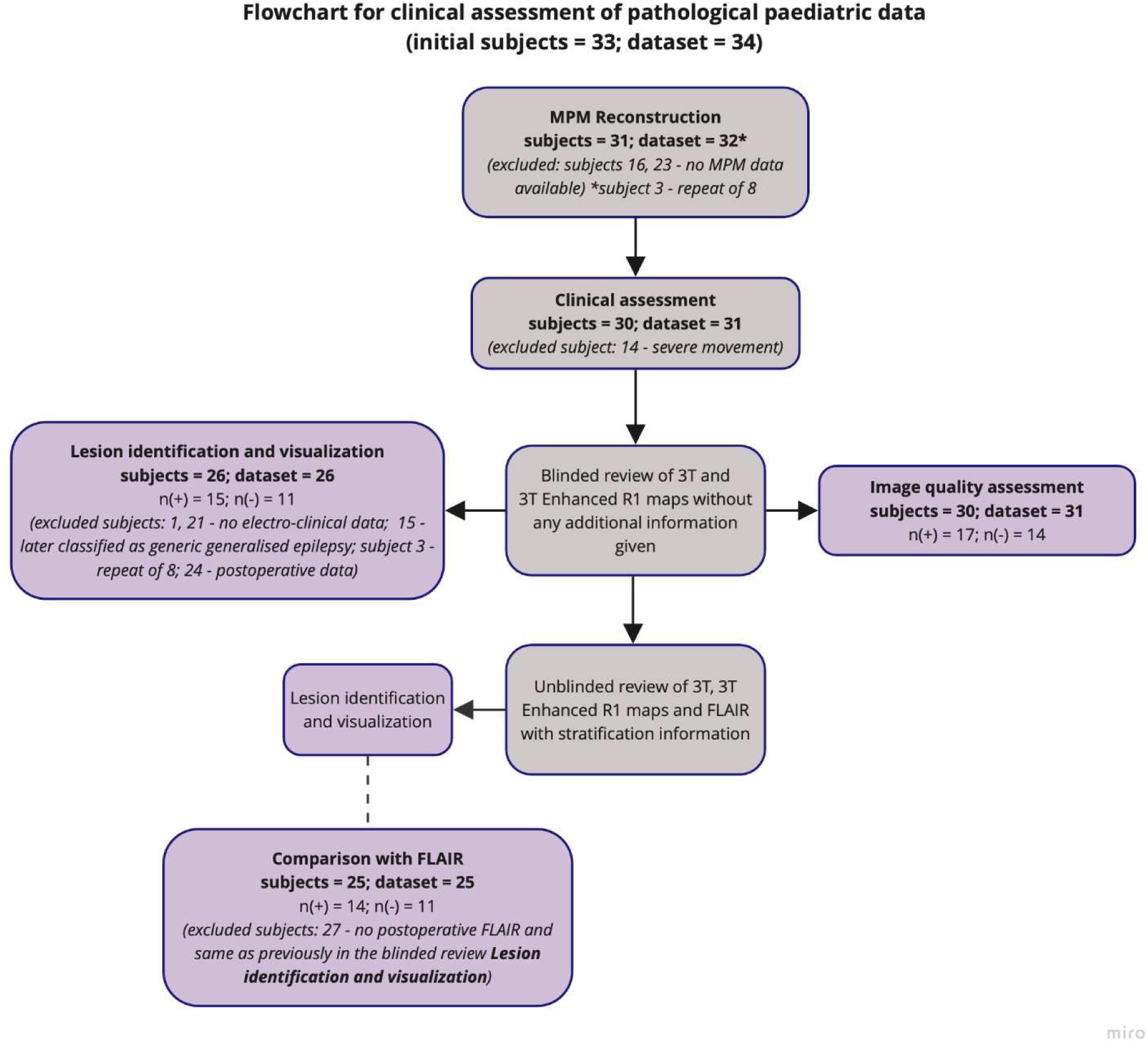
The flowchart includes the initial number of participants, those excluded for any reason, as well as the final numbers used for each analysis.

**Table 1:**
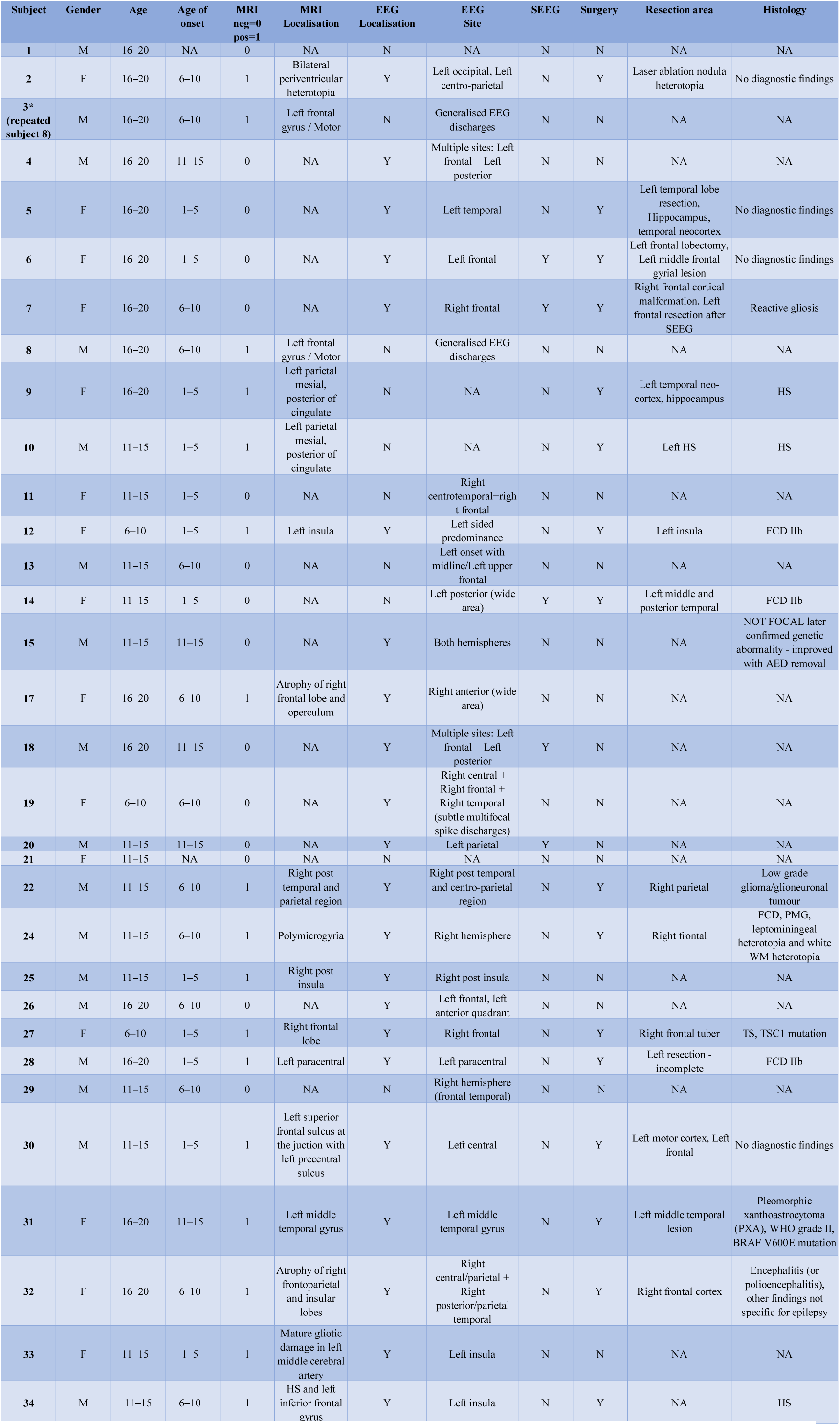
Demographic and clinical data of the pathological paediatric population.

#### IQT Data analysis

Thirty-one R1 maps were enhanced applying the hyper-parameters and weights derived from the training on DataMPI to reconstruct AI Enhanced 3T R1 maps.

#### Image quality assessment: 3T versus AI Enhanced R1 maps

In a first review step, 3T and AI enhanced R1 maps were presented to a paediatric consultant neuro-radiologist with 10 years of experience (FD’A) in a randomized order blinded to any other clinical information including their status as MR-positive/negative. Images were assessed for GM-WM sharpness of the cerebrum, the cerebellum and the basal ganglia using a scale of 1-4 (1 (not visible), 2 (subtle), 3 (clear) or 4 (very clear)). The ranking results were statistically analysed using a non-parametric Wilcoxon test for 2 related samples in SPSS and frequency of the scores bar chart was generated. Significant differences were based on a threshold of p<0.05.

#### Epilepsy lesion identification: 3T versus AI Enhanced R1 maps

Next, both 3T and AI enhanced R1 maps were then reviewed for lesions blinded to any other electroclinical information. Lesion visibility was also scored using a scale of 1-4 (1 (not visible), 2 (subtle), 3 (clear) or 4 (very clear)).

#### Epilepsy lesion identification: Comparison to FLAIR

In a final review step 3T and AI enhanced R1 maps were presented together in a randomized patient order, unblinded to stratification information (MR positive/MR negative, MR localization if MR positive and EEG localization with a detailed description of the electro-clinical findings, including the suspected hemisphere and lobe). At this stage the maps were reviewed together with their clinical 3T 3D FLAIR[23](typically the best scan for lesion detection in epilepsy MRI protocols[24,25]) for comparison purposes. Lesions were scored for visibility using the scale described above for each image type.

Following this review process the number of lesions detected in the blinded and unblinded review were summarized in the MR positive and negative groups. Lesion visibility scores were also compared between the 3T and AI enhanced R1 maps and 3D FLAIR.

## Results

Enhanced R1 maps were obtained in all of the 31 cases scanned. Enhanced images were of high quality and could be seen to contain pathological abnormalities present on the 3T maps that were not present in the healthy control adults training dataset (*Figure 2*).

**Fig. 2:**
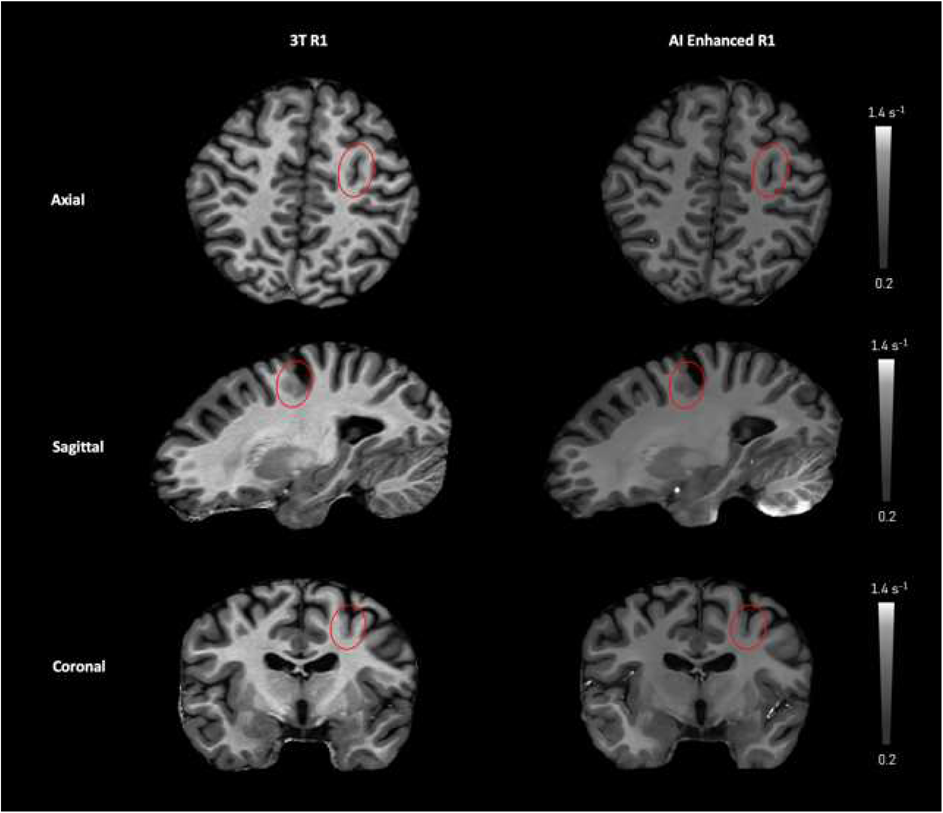
Lesion identified in subject 8 in axial, sagittal and coronal view for the 3T (left) and AI Enhanced (right) R1 maps. Note images are scaled to quantitative units (s^-1^) and R1 values are smaller at 7T than 3T so maps appear darker.

### Image quality assessment: 3T versus AI Enhanced R1 maps

AI enhanced R1-maps had superior image quality compared to the original 3T R1-maps (*Figure 3*, *table 2*). The Wilcoxon Signed Ranks test showed 31 positive ranks (AI Enhanced > 3T), no negative ranks (AI Enhanced < 3T) and no ties (AI Enhanced = 3T) for the cerebrum and the basal ganglia, and 27 positives, 4 negatives and no ties for the cerebellum.

**Fig. 3:**
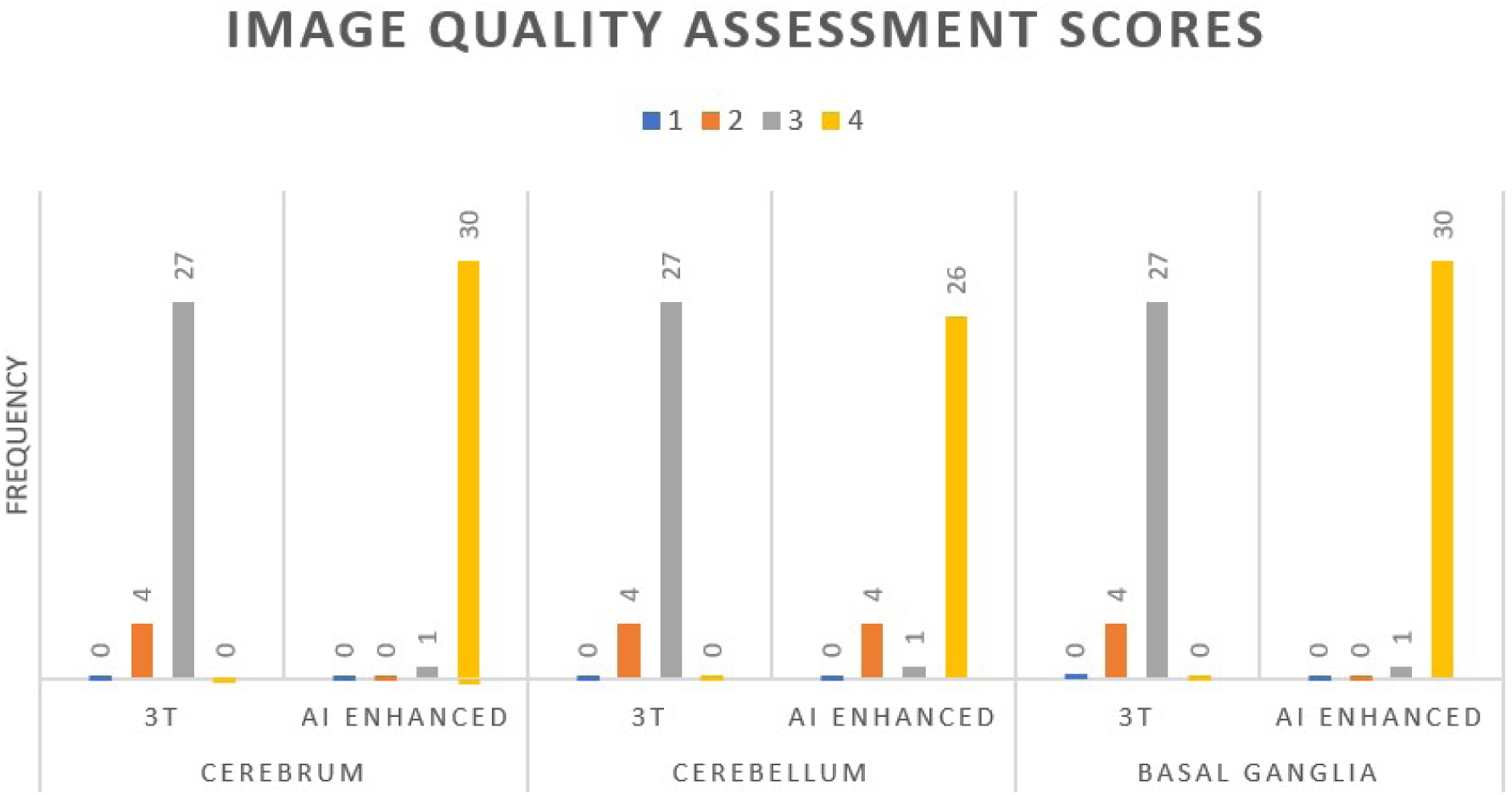
Bar chart for the radiological image quality assessment scores frequency (y axis) for the 3T and the 3T Enhanced R1 maps in the cerebrum, cerebellum and basal ganglia. 1-4 (1 (not visible), 2 (subtle), 3 (clear) or 4 (very clear)).

**Table 2:** Image Quality Assessment Scores and Their Interpretations.

| Image Quality Assesment Score | Interpretation |
| --- | --- |
| 1 | not visible |
| 2 | subtle |
| 3 | clear |
| 4 | very clear |

### Epilepsy lesion identification in the MRI positive cohort

#### 3T versus AI Enhanced R1 maps

In the radiological review blinded to any additional clinical information, 3T R1 maps had lesions detected in 9 out of the 15 MRI positive cases, while 11/15 lesions were detected in the AI Enhanced R1 maps. The lesion visualization was scored better in 9/11 of the AI Enhanced maps compared to the 3T maps and equivalent in the remaining two cases. In the unblinded radiological review the identified lesion numbers increased to 13/15 and 14/15 for the 3T and AI Enhanced R1 maps, respectively.

#### Comparison to 3D-FLAIR

Lesion visualization was assessed in 14 MR positive subjects comparing the 3T R1 maps, AI enhanced maps to clinical 3D-FLAIR unblinded (with additional electroclinical information provided). The visualization of lesions was better in the AI Enhanced R1map compared to the 3D-FLAIR in 6/14 (see examples in *fig. 4A* and *4B*) (1 out of these 6 was judged as ‘not visible’ on the 3D-FLAIR *fig. 4C*); 5/14 were the same and in the remaining 3/14 visualisation was judged as worse than the 3D-FLAIR (in 1 out of these 3 was judged as ‘not visible’ on AI Enhanced). The visualization of lesions was worse on the 3T R1 maps compared to 3D-FLAIR. In 2/14 it was better (1 of 2 was judged as ‘not visible’ on FLAIR), 7/14 the same and in 5/14 it was worse (of which two were ‘not visible’).

**Fig. 4:**
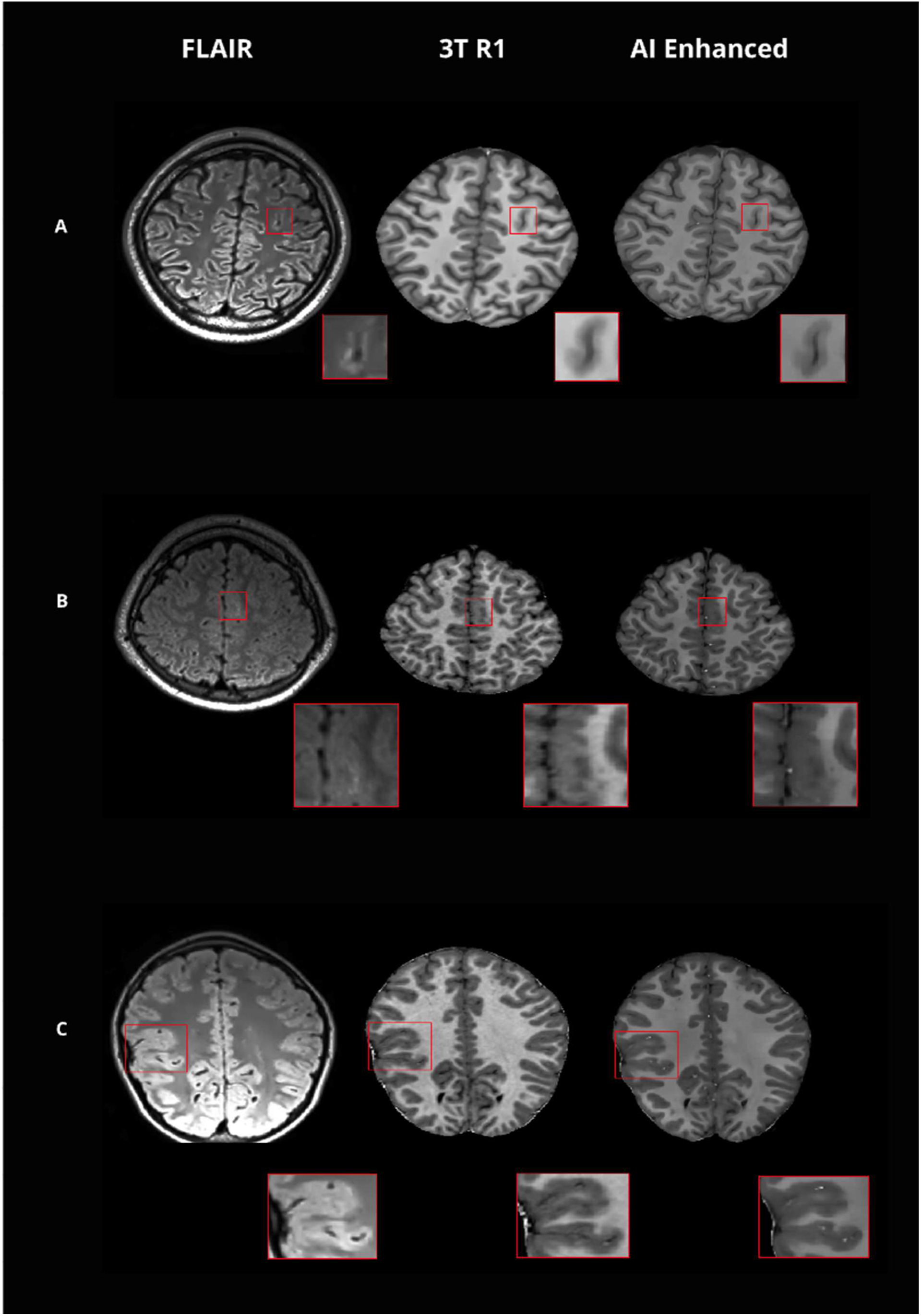
*Examples of MR positive cases (**A** (**subject 8), B** (**subject 28**)), where the visualization of the lesion with the 3T Enhanced was superior to the 3T R1 and FLAIR. Example of MR positive case (**C** (**subject 10**)), where the lesion visualization was characterized as: FLAIR ‘negative’, 3T ‘subtle’, 3T Enhanced ‘clear’*.

#### Epilepsy lesion identification in the MRI negative cohort

In the blinded radiological review in the MRI negative cohort, lesions were not detected in the 3T R1 maps, however, in 4/11 cases lesions were identified in the AI Enhanced R1 maps (subjects 7, 13, 26, 29).

#### Comparison to 3D-FLAIR

Unblinded review of R1 maps and comparison of AI enhanced R1 maps to 3D-FLAIR (*table 3* green): Seven out of 11 had no lesion identified on the 3D-FLAIR nor the R1 maps (1 of these 7 (subject 18) was noted as suspicious for transmantle sign in left mesial frontal on the AI Enhanced map). For the 4 cases with lesions identified, in 3 the AI enhanced map was superior to the FLAIR which was scored as ‘not visible’ as was the 3T R1 map. However, the AI Enhanced maps were scored as having ‘subtle’ lesions (*fig. 5A* (subject 7), 5B (subject 13) and 5C (subject 26)). In the remaining MR negative case (*fig. 5D* (subject 29)) where a lesion was detected both the 3D-FLAIR and AI enhanced maps were scored as having a ‘subtle’ lesion while the 3T R1 map was ‘negative’.

**Fig. 5.**
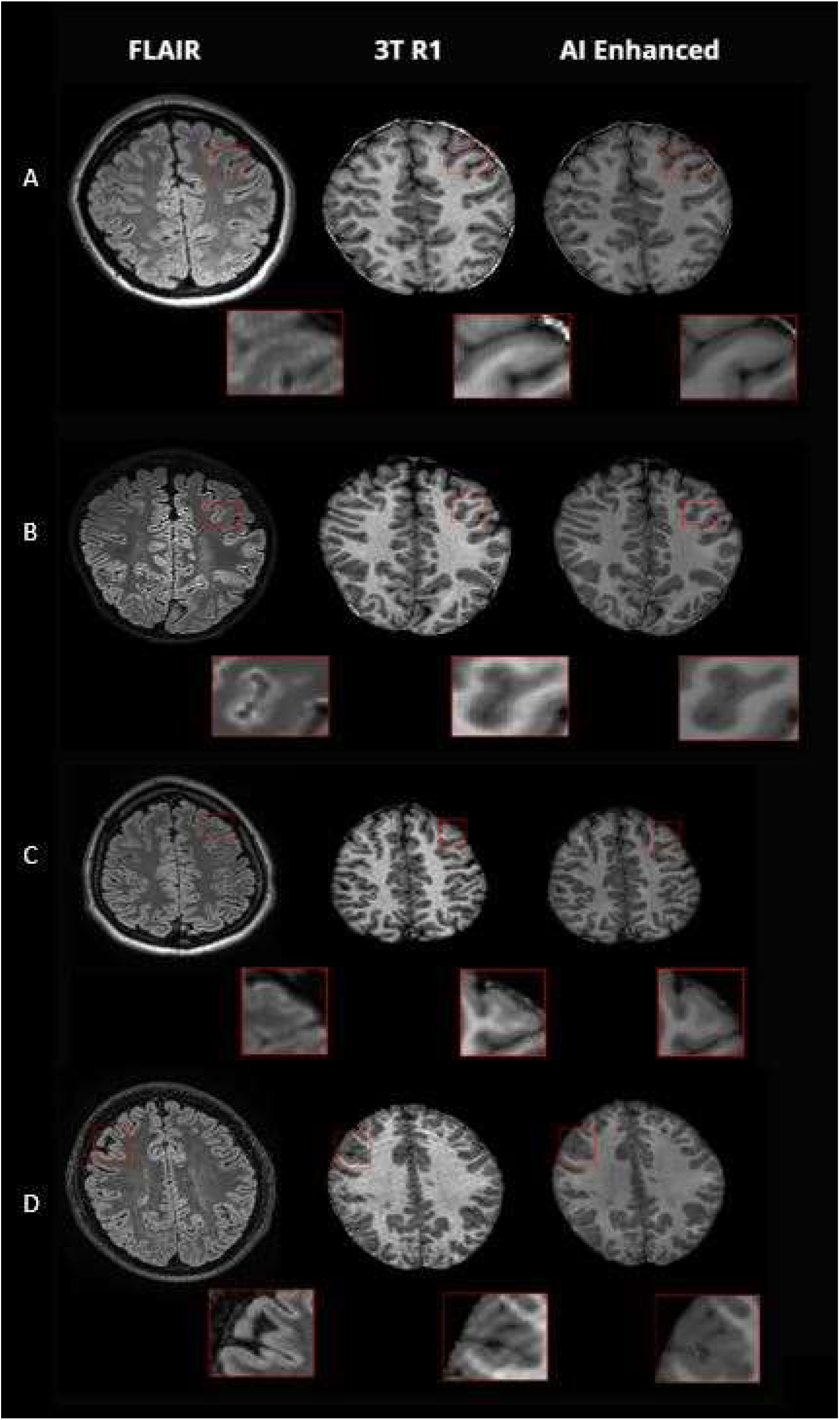
**A-C** MR negative cases with lesions identified only on the AI Enhanced R1 map (**A** (**subject 7), B** (**subject 13**), **C** (**subject 26**)). **D** (**subject 29**) the MR negative subject with a lesion identified on both the AI Enhanced R1 map and 3D FLAIR.

**Table 3:**
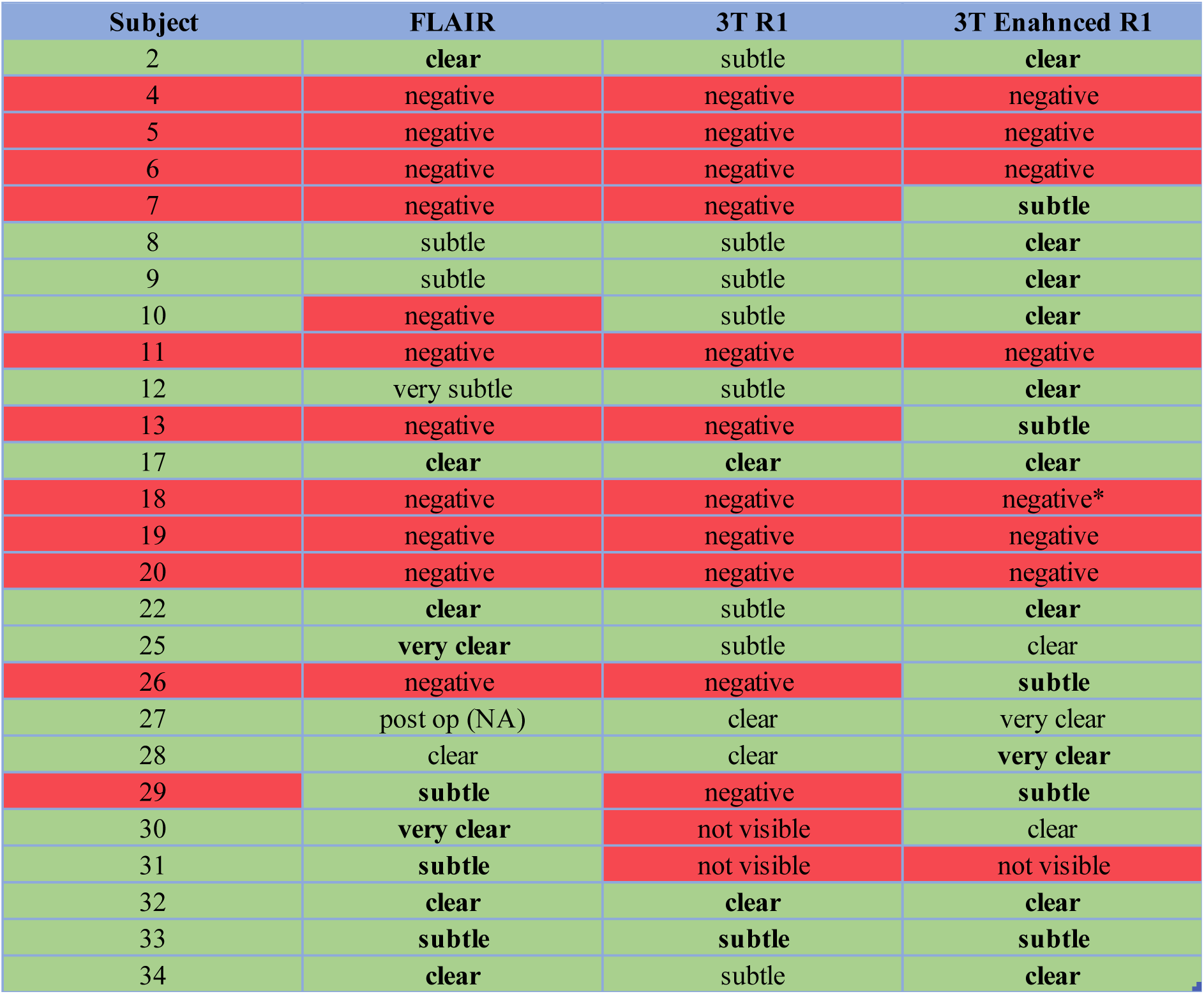
Clinical assessment – lesion identification results for unblinded review. Column ‘Subject’ presents the subject’s stratification (red for no lesion seen and green for lesion identified) based on clinical 3T epilepsy protocol and neuroradiology expert review at the time where the MPM data were acquired; Similarly, ‘FLAIR’, ‘3T R1’ and ‘AI Enhanced R1’ columns show whether there was a lesion identified after the radiological score of how visible this lesion was on a 4 point Likert scale (not visible, subtle, clear,

In these 4 MR negative cases, Subject 7 had a left frontal lesion detected which was concordant with SEEG and resection. Follow up 3T clinical MRI remained negative. Subject 13 had a left frontal lesion detected which was concordant with scalp EEG and a follow up repeat clinical 3T MRI. In subjects 26 and 29, lesions were concordant with EEG and seizure semiology, but no further investigations were performed to confirm these results.

## Discussion

We aimed to improve image quality by creating high resolution AI enhanced R1 maps from lower resolution 3T R1 maps using a U-Net patch-based IQT network trained on data from a group of ten healthy individuals scanned at both 3T (1mm) and 7T (0.5mm). AI enhanced R1 maps were then generated in a cohort of paediatric epilepsy patients (separate scanner and site), exploiting the cross-site consistency of quantitative maps[10,26]. The clinical impact of the AI enhancement was evaluated on this paediatric epilepsy cohort consisting of both MR positive and MR negative patients with electro-clinical features consistent with MCD.

We firstly determined if the AI enhanced R1 maps had improved image quality, and that the trained network could generalize to the test dataset. Radiological assessment of image quality in the cerebrum, basal ganglia and cerebellum showed that the AI enhanced images scored significantly higher than the 3T R1 maps (*fig. 3*). We then evaluated, if imaging abnormalities were faithfully reconstructed in the AI enhanced images. In the 14 of 15 paediatric patients with visible lesions (MR-positive cohort) the enhanced images were able to preserve the pathological features even though there were no pathological cases in the training dataset of healthy adults (see examples in *fig. 4*). In the remaining subject the lesion identified in the clinical report was not confirmed by our expert rater (FD’A) on 3D-FLAIR.

Secondly, we determined if lesion visibility was enhanced in MR positive cases and if the enhanced R1 maps might increase the yield of lesion detection in MR negative cases. The lesion identification and visualization assessment within the MR positive cohort showed that the AI Enhanced R1 maps were always either superior (11/15) or equivalent to (4/15) the 3T R1 maps. The AI enhanced R1 maps enabled the expert reviewer to identify 4 new lesions out of the 11 MR-negative patients (the newly detected abnormalities were not visible in the original 3T R1 maps in all cases and were not visible in the 3D-FLAIR in 3/4). In one further case the AI enhanced map showed some indication of a transmantle sign, an abnormality associated with FCD IIb but it was not judged clear enough to be classified as a lesion.

Finally, we compared the lesion visibility in the maps to the clinical ‘gold standard’ 3T 3D-FLAIR image. The AI enhanced images had improved visibility in 6/14, similar in 5/14 and worse in 3/14. In the MR negative cohort 4 of 11 lesions were identified on the AI Enhanced R1 maps; only 1 of these 4 was also judged to be ‘subtle’ on the 3D-FLAIR. Validation of these new results in the four MR negative patients is challenging because relatively few of these patients go on to have surgery. The localisation of 1 of 4 lesions identified on the 3T Enhanced was concordant with SEEG findings and resection area with ILAE I seizure outcome following surgical resection (subject 7). In a further subject the localisation was concordant with EEG and a subsequent repeat clinical MRI (subject 13). In the remaining two MR negative patients with new findings the abnormalities were concordant with EEG but they did not have further investigation with SEEG or resective surgery (subject 26 and 29).

Some previous studies have utilised 3T T1 or R1 maps to detect abnormalities in FCD[3,27,28] although limited proven benefit for lesion detection in MRI negative cases was found[29]. Increased T1 values are correlated with baseline myelination and susceptible to microstructural damage likely to disrupted myeloarchitecture[30]. Here, we utilised R1 maps primarily to allow for IQT network training on 3T and 7T data from one site to be used on paediatric data from another, leveraging the high cross-site reproducibility of quantitative methods[10] that remove hardware specific influences. IQT, being patched based, was then able to generalise to the new data despite a small training dataset from 10 healthy individuals scanned at both field strengths.

Multiple features contributed to the improved radiological visualisation; in addition to increased resolution, the AI enhanced maps had reduced noise and motion artefacts that were not in the training data (*Fig 6*). Training data limitations were transferred to the enhanced maps including bright vessels owing to the 7T head transmit coil and low B1 transmit fields in the cerebellum that could be mitigated using parallel transmit approaches[31].

**Fig. 6:**
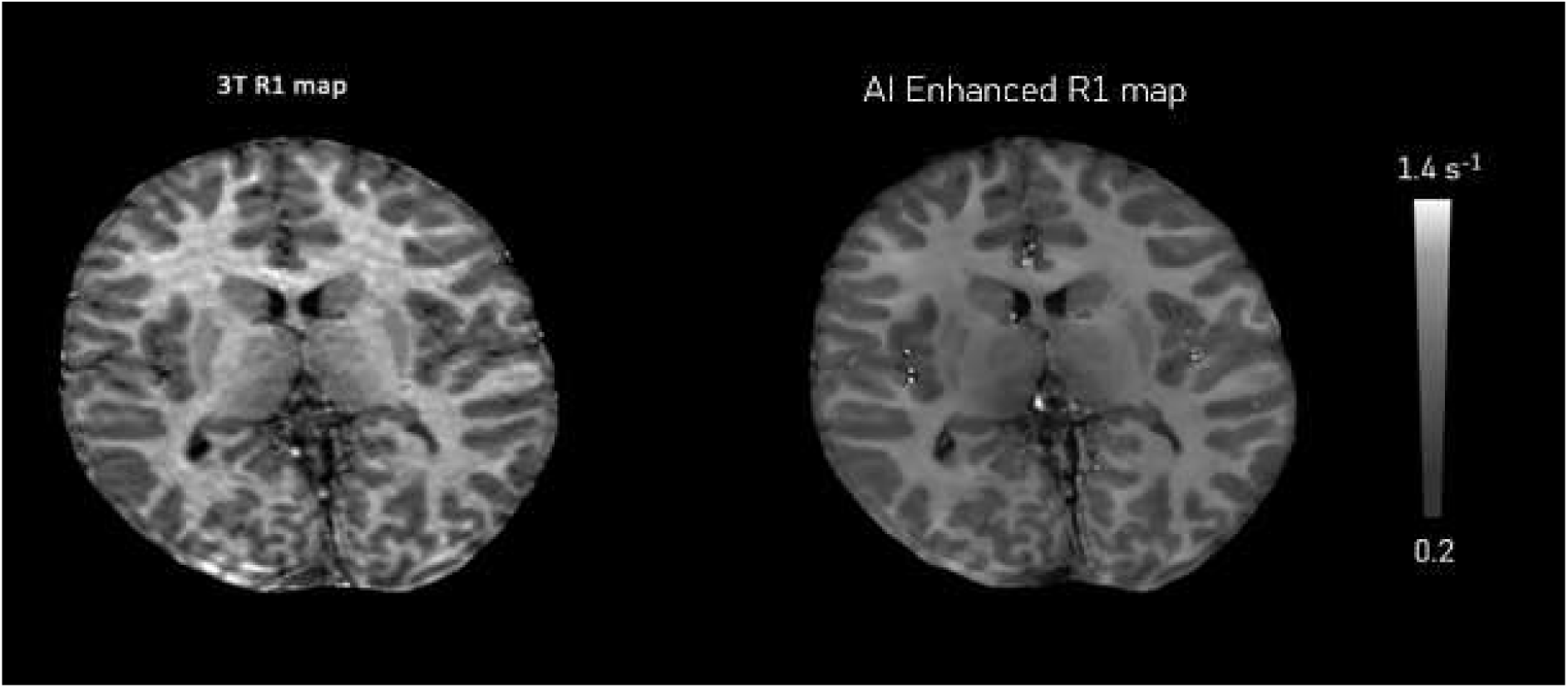
Example of a 3T R1 map which suffers from motion artefact and the image quality has been improved on its 3T enhanced version.

Using AI to enhance 3T R1 maps showed a yield of 14/15 (93%) lesions in the MR positive cohort, in the remaining case the clinically reported lesion was not confirmed by our expert in any scans. In 4/11 (36%), MR negative patients, lesions were detected, a similar yield to direct 7T imaging studies with ranges from 17-44%[4,32–34]. Interestingly, 3 out of 4 were consistent with electro-clinical information but judged not visible on clinical 3T 3D-FLAIR images even when guided by the AI enhanced R1 map; none were detectable on the 3T R1 maps highlighting that a genuine and clinically relevant image quality improvement had been achieved. The ability to enhance 3T data has advantages over UHF imaging owing to reduced cost and increased availability. Limited validation of new findings was possible owing to the rarity of surgery and SEEG in MR negative subjects that requires a larger prospective study. It would be possible to enhance 3D-FLAIR images, however signal features related to imaging parameters and hardware reduce generalisability across sites. While focal epilepsy was the application here, the approach is readily applicable to a wide range of potential patient groups where enhanced image quality could impact patient management.

## Abbreviations

SNR: Signal-to-Noise Ratio
CNN: Convolutional Neural Networks
IQT: Image Quality Transfer
MCDs: Malformations of Cortical Development
FCD: Focal Cortical Dysplasia
MRI: Magnetic Resonance Imaging
GM: Gray Matter
WM: White Matter
MPM: Multi-Parameter Mapping (MPM)
HS: Hippocampal Sclerosis

## Data Availability

All data, and materials are available on request to any researcher subject to any ethical or data protection restrictions. All code is freely available for any research, non-commercial purpose.

## Acknowledgments

Thanks for the assistance of Dr Peter McColgan and Dr Kerrin Pine for their assistance in the training data collation.

## Author contributions

Conceptualization: DWC, DCA

Methodology: GD, DCA, DWC, MF, HL, FD’A

Investigation: SL, GD, RP

Visualization: SFB, MJM, JLS, EH

Funding acquisition: DWC, HJC, DCA

Project administration: DWC, DCA

Supervision: SJ DWC, DCA, JOM

Writing – original draft: GCA, DWC

Writing – review & editing: GD, FD’A, MF, HL, SL, RP, JOM, JHC, NW, DCA, DWC

## Data and materials availability

Thanks for the assistance of Dr P McC and Dr K P for their assistance in the training data collation.

